# Heart rate variability at ICU admission in Covid-19 patients in sitting position: a prospective study

**DOI:** 10.64898/2025.12.25.25343013

**Authors:** Tomás Francisco Fariña-González, Fernando Martinez-Sagasti, María Elena Hernando, Ignacio Oropesa, Antonio Nuñez-Reiz, Miguel Ángel González-Gallego, Julieta Latorre, Manuel Quintana-Diaz

**Author notes:** **Corresponding author:** Tomás Francisco FARIÑA-GONZÁLEZ.

## Abstract

**Purpose:** to describe HRV metrics in Covid-19 patients at admission in the ICU and its relationship with mortality and invasive mechanical ventilation (IMV). Heart rate variability (HRV) in sitting position in critically ill patients has not been explored.

**Methods:** we conducted a prospective single-centre observational study. Adult patients admitted in the ICU with respiratory failure due to RT-PCR-confirmed SARS-CoV-2 but not under IMV were included. Electrocardiogram was recorded at least for 15 minutes at 500 Hz during a stable sitting condition. Power spectrum was obtained using wavelets. Very low frequency (VLF), low frequency (LF) and high frequency (HF) powerbands were calculated and then normalized to total power (VLFn%, LFn% and HFn%). We also analyzed non-linear HRV dynamics.

**Results:** 27 patients were included. LFn% was lower in non-survivors (4.5 vs 8 %, p=0.015) and related to 28-day mortality (OR 0.61; 95% CI: 0.32 to 0.9, P=0.05). In a robust generalized Gamma linear model, we found that detrended fluctuation analysis alpha 2 (DFA⍺2) (RR 0.092; bootstrapped 95% CI: 0.031-0.361, P=0.003), admission APACHE II score (RR 1.084; bootstrapped 95% CI: 1.046 to 1.123, P=0.002) and SAF index (RR 0.985; bootstrapped 95% CI: 0.982 – 0.990, P<0.001) were associated with longer ICU LOS.

**Conclusions:** diminished normalized LF power was associated with 28-day mortality in univariate analysis among critically ill COVID-19 patients on spontaneous ventilation, potentially reflecting an altered autonomic response in early severe COVID-19. Normalized and absolute VLF power should be considered when analyzing HRV in ICU patients. DFA⍺2 was the HRV variable with the strongest association with ICU LOS. These exploratory results could be helpful to design newer tools for early prognostication in COVID-19 patients.

## BACKGROUND

SARS-CoV-2 (also known as Covid-19) is a coronavirus discovered in December 2019 in China which unleashed one of the worst pandemics ever known to humanity (1, 2). Although the main clinical features are related to respiratory disease, other organs are also directly affected by the virus. Myocarditis, renal injury and skin involvement have been described (3–5). Neurological symptoms and syndromes are also found in the literature (6, 7). Neurological effects of Covid-19 are multiple, some related to the primary viral injury but others are due to the immune response or administered therapies. These neurological factors affect the central and peripheral nervous system (8, 9).

Interestingly, many patients present a clinical dissociation between hypoxemia and their cardiovascular response; tachycardia or hypotension are not usually found in contrast to other severe infections. Also, hypoxemia is disproportional to the dyspnoea referred by the patients, a situation called “happy hypoxemia” (10, 11). These characteristic features might be related to the viruś neurotropism (12). SARS-CoV-2 has been found in the carotid body (13), an important centre in regulation of reflex changes in ventilatory and autonomous responses (14).

Heart rate variability (HRV) is an intrinsic property of cardiovascular dynamics, describing changes of the intervals between consecutive beats. This cardiovascular response to stress (such as hypoxemia) is mediated by the autonomic nervous system (ANS) and HRV is an important part of it. Automaticity is a main feature of cardiac pacemaker structures; however, heart rate and rhythm are tightly controlled by the ANS in order to match tissue metabolic demands to the oxygen delivery (15). Therefore, HRV has been proposed as a surrogate measure of this cardiac autonomic control and has been studied in many different diseases (16–25). Early mobilization results in better outcomes in critically ill patients and is advocated in actual guidelines to be good practice, if feasible (26, 27). Kloss et al., in a multicenter study supported by the European Society of Intensive Care Medicine, found that those surviving Covid-19 patients who were mobilized early within 72 h of ICU admission (i.e. to a sitting position) had an increased chance of being discharged home (28). Body position modifies HRV metrics (29–32), however we found no previously published studies exploring sitting position in critically ill patients.

HRV can be described using several methods. Assuming stationarity, the investigator can use statistical and geometric metrics (time domain) or decompose RR intervals in their primary frequencies (frequency domain) to analyse the signal. Although easy to compute, they are prone to error if artifacts (such as ectopic beats) are introduced in the signal. Mostly published studies about HRV in the ICU setting use short-term (5-minute) records, following the 1996 Task Force guidelines (33). This approach correctly evaluates low (LF) and high frequency (HF) events, however very low frequency (VLF) events, those that happen between 0.003 Hz to 0.04 Hz, might not be clearly described as they might happen once every 5 minutes. Moreover, biosignals are dynamic due to complex interactions between different systems and, therefore, are not stationary. Non-linear methods, such as Poincaré plots, detrended fluctuation analysis, sample entropy or power spectral index overcome these issues (20). HRV has been integrated in predictive models for critically ill patients, particularly for early identification of complications or even death (34–39).

## METHODS

Our main objective was to describe HRV metrics in Covid-19 patients in sitting position in the first 24 hours of admission in the ICU and to explore if it can be feasible to predict 28-day mortality and invasive mechanical ventilation need.

We performed a prospective single-centre observational study. Adult patients admitted to our 46-bed Department of Critical Care at Hospital Clínico San Carlos from 18/03/2021 to 31/12/2021 with respiratory failure due to RT-PCR-confirmed SARS-CoV-2 but not under IMV were included during the morning shift. Exclusion criteria were previous history of diabetic neuropathy, history of neurologic autonomic disease, atrial fibrillation and recent (< 3 months) brain trauma. We also excluded those patients who could not be followed during the study period (14 days from ICU admission).

Data and blood samples for biochemical analysis were collected within 24 hours from admission in spontaneous ventilation. Each patient was connected to a Phillips IntelliVue™ monitor system. A laptop was connected to the monitor to synchronously download the waveforms by means of a dedicated open source software (40). Waveforms were continuously recorded at least for 15 minutes at 500 Hz during a stable sitting condition, in the morning time. All recordings were made by the same investigator (TFF) who did not interfere in clinical decisions. We analysed EKG raw data, extracting RR intervals using HRVtool package for MatLab (41). Each recording was visually inspected for accurate R wave detection. Artifacts and irregular beats were manually deleted. RR peak times were then exported to R and analysed with the RHRV package (42). We interpolated the input data at 6 Hz, using a linear approach. The study protocol was approved by the local Ethics committee (CI 21/169.E) prior to the beginning of the trial and first patient enrollment and followed the regulations established by the Helsinki Declaration. Written informed consent was obtained from all patients or their next-of-kin / guardians.

HRV METRICS: EKGs are non-stationary signals. Therefore, for power spectrum analysis we used the wavelet analysis approach using extremal phase Daubechies d4, based in the Maximal Overlap Discrete Wavelet Packet Transform (MODWPT) (43). Spectral indices obtained from power spectra included very low frequency (VLF, 0.003-0.04 Hz) low frequency (LF, 0.04–0.15 Hz) and high frequency (HF, 0.15-0.4 Hz). Also, total power, which represents the total area under the spectrum and a measure of the overall variability of the series, and the relative power of VLF, LF and HF powerbands (VLFn%, LFn% and HFn%) were calculated. We also analyzed non-linear dynamics with Poincaré plots, detrended fluctuation analysis and correlation dimension (20, 44).

STATISTICAL ANALYSIS: In a multicenter, prospective observational study in 50 countries, Bellani et al. (45) described an ICU mortality of 35.3% in no-Covid acute respiratory distress syndrome. About Covid-19 patients, we found in our records (unpublished) that approximately 50% of the patients admitted to the ICU required IMV in the “second” and “third” waves (June 2020 to December 2020) of the Covid-19 pandemic. In a similar population to ours, Aragon-Benedí et al. found 51% ICU mortality rate in patients with Covid-19 (46). To calculate sample size, we considered median LF/HF ratio values in ICU patients published in the systematic review by Karmali et al (47). Considering a LF/HF value 2±0.5 in ICU patients and 30% difference in our population, for a statistical power of 80% (α 0.05, β 0.2), a sample size of 22 patients was calculated.

For analysis, patients were separated in two mortality-groups (survivor/non-survivor) and then compared using non-parametrical tests (Mann-Whitney U test). Continuous variables are presented as median with the interquartile range (IQR) and number of patients (percentage) for qualitative data. p <0.05 was considered significant. Due to the small sample size and the presence of outlier values that might affect the robustness of analyses like binomial logistic regression or Ordinary Least Squares linear regression, ad hoc removal of records or variable transformation were not considered appropriate. In order to maintain statistical rigor, robust methods were used for the analysis. Multivariate regression models were built using two or three predictors, those with the lowest p values and with clinical interest, to minimize overfitting risk and maintain an adequate events-per-variable ratio. •Firth’s logistic regression was performed for 28-day mortality each variable and those with a potential influence (p<0.1) were introduced in a multivariate analysis. A robust Gamma generalized linear model was performed to describe ICU LOS predictors. Bootstrapping (2000 iterations) was used to calculate p values, rate ratios and confidence intervals. When comparing results against previously published data, we used one-sample Wilcoxon signed-rank test. Rstudio v. 2024.12.1+563 (Posit Software, USA) and R packages: robustbase, ggplot2, broom, dplyr, cowplot, boot, MASS, tidyr and logistf were used for statistical analysis.

## RESULTS

Forty-seven patients were screened, 29 patients did not meet the inclusion criteria, and one was excluded did not speak Spanish and thus unable to give consent. Therefore, 27 patients were finally recruited (Fig 1).

**Fig 1.**
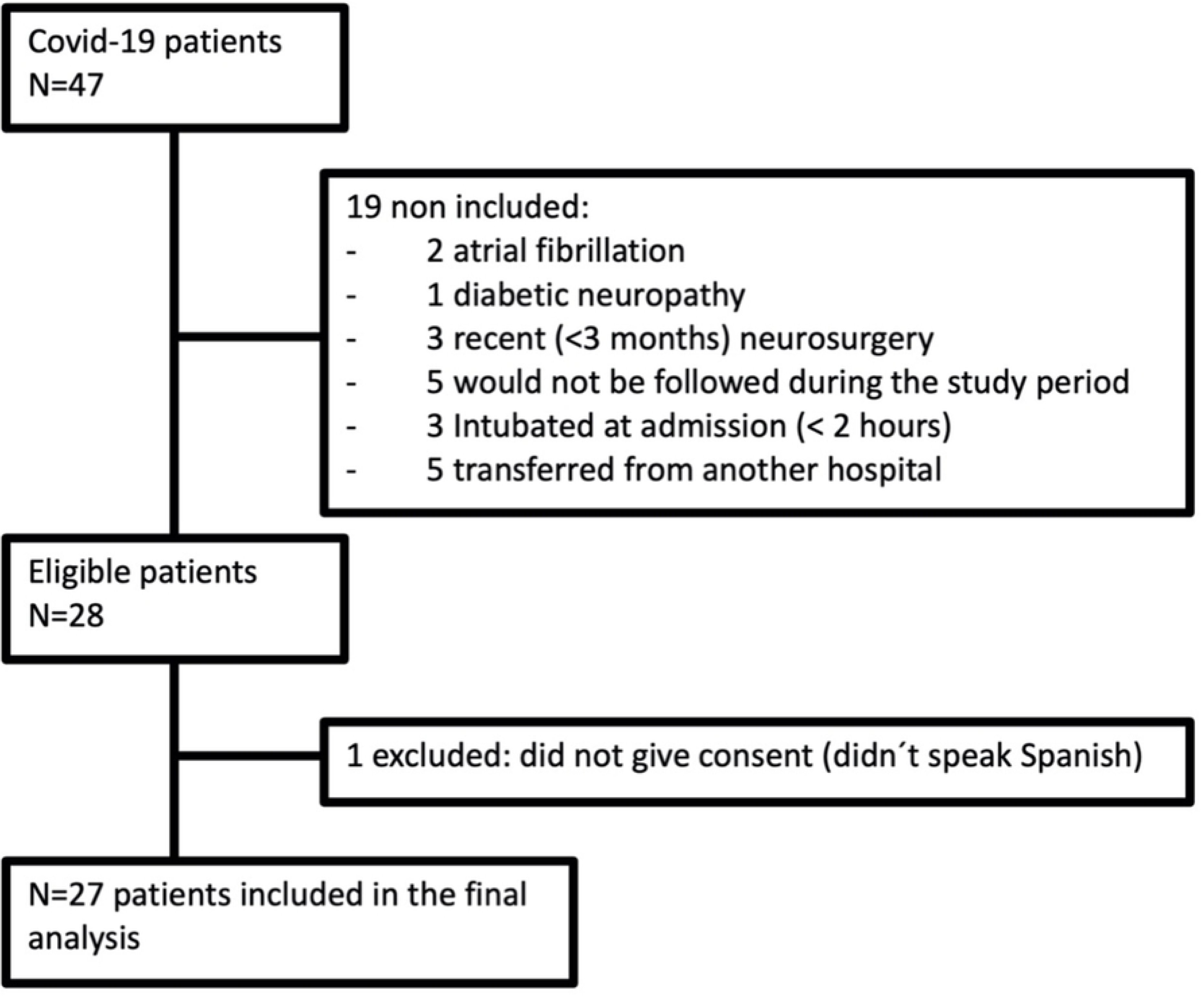
Flow chart

Out of the 27 patients included, 6 patients (22.2%) were dead at day 28. Non-survivors were mostly male, 5 patients (83.3%), and older (73 vs 59 years, p=0.036) than survivors. All patients who did not survive had arterial hypertension (100 vs 23.8 %, p=0.002). Renin-angiotensin-aldosterone system inhibitors (RAASi) were the most usual treatment (100 vs 23.8%, p=0.008). Interestingly, betablockers were not commonly used in these patients (16.7 vs 9.5 %, p=0.545).

There were no differences in time of hospital and ICU admission. APACHE II score was higher in non-survivors but this difference was not statistically significant. Although median ICU (ICU LOS) and hospital length of stay were higher in non-survivors, there was no statistical significance.

All non-survivors were intubated. Renal replacement therapies were more common in non-survivors (66.7 vs 19%, p=0.044). The median from ICU admission to intubation was 3.5 days (IQR 2-8) in non-survivors and 6 days (IQR 3-8) in survivors (table 1).

**Table 1.** Demographics and background for 28-day mortality groups.

|  | No survivor at day 28 (n=6) | Survivor at day 28 (n=21) | p |
| --- | --- | --- | --- |
| Male gender, n (%) | 5 (83.3) | 14 (66.7) | 0.633 |
| ***Age years, median (IQR)*** | ***73 (63.25-74.75)*** | ***59 (37-69.5)*** | ***0.036*** |
| Body mass index, median (IQR) | 27.25 (25.57-31.41) | 26.9 (25.15-31.3) | 0.842 |
| APACHE II score, median (IQR) | 14 (11-18.25) | 9 (8-14.5) | 0.125 |
| Diabetes, n (%) | 2 (33.3) | 5 (23.8) | 0.633 |
| ***Arterial hypertension, n (%)*** | ***6 (100)*** | ***5 (23.8)*** | ***0.002*** |
| Betablockers, n (%) | 1 (16.7) | 2 (9.5) | 0.545 |
| ***RAASi, n (%)*** | ***5 (83.3)*** | ***4 (19)*** | ***0.008*** |
| Days from symptoms to hospital admission, median (IQR) | 5.5 (0.75-7) | 7 (3.5-9) | 0.14 |
| Days from hospital admission to ICU, median (IQR) | 1 (0.75-2.75) | 2 (1-3) | 0.476 |
| Antibiotics at day 1, n (%) | 2 (33.3) | 5 (23.8) | 0.633 |
| Corticoids at day 1, n (%) | 6 (100) | 20 (95.2) | 0.778 |
| ***Intubated, n (%)*** | ***6 (100)*** | ***9 (42.9)*** | ***0.02*** |
| ***Need of RRT, n (%)*** | ***4 (66.7)*** | ***4 (19)*** | ***0.044*** |
| LOS ICU, days median (IQR) | 25.5 (19.25-36.25) | 14 (7.5-41) | 0.316 |
| LOS Hospital, days median (IQR) | 25.5 (19.25-36.25) | 21(16-45.5) | 0.887 |
| Time to intubation, days median (IQR) | 3.5 (0.75-7) | 6 (3-8) | 0.272 |

Comparisons using Fischer's exact two-tailed test for dichotomic variables and Mann-Whiney U test for continuous variables. In bold and italics, $p < 0.05$ . RRT, renal replacement therapies. RAASi, Renin-angiotensin-aldosterone system inhibitors. LOS: length of stay

### CLINICAL AND BIOCHEMICAL PARAMETERS

Non-survivors were slightly more tachypneic than survivors, although not reaching statistical significance (28.5 vs 25 breath/min, p 0.316). There were no differences in oxygenation variables as described by the ROX index (5.02 vs 6.31, p 0.242) and SAF index (111 vs 145, p 0.457).

There were no differences in cardiac biomarkers troponin I and ProBNP (0.013 vs 0.1 ng/L and 316 vs 259 pg/mL, p=0.195 and 0.469, respectively).

Median IL-6 and procalcitonin were higher in non-survivors (67.8 vs 21 pg/mL and 0.28 vs 0.11 ng/mL, p=0.03 and 0.049) but C-reactive protein and proadrenomedullin were not different (median 20 vs 8.54 mg/dL and 1.06 vs 0.8 nmol/L, p=0.589 and 0.199).

### HRV ANALYSIS (table 2)

Recordings lasted a median of 36 minutes in non-survivors versus 41 minutes in survivors. There was no difference in average heart rate (78.76 beats/min in non-survivors vs 80.4 beats/min in survivors, p=0.755). Median RR interval was also similar.

**Table 2.** Heart rate variability (HRV) metrics according to 28-day mortality.

|  |  | Non survivor at day 28 (n=6)<br>Median (IQR) | Survivor at day 28 (n=21)<br>Median (IQR) | p |
| --- | --- | --- | --- | --- |
|  | Mean RR interval msec | 762 (706.5-859.25) | 749 (679-877) | 0.755 |
|  | Mean HR<sub>60</sub> | 78.76 (70.14-85.2) | 80.4 (68.43-88.41) | 0.755 |
| Wavelet | VLF power msec<sup>2</sup> | 599.89 (377.87-1359.89) | 1086.14 (393.64-1796.61) | 0.629 |
|  | LF power msec<sup>2</sup> | 28.62 (13.1-86.6) | 70.43 (40.91-169.72) | 0.097 |
|  | HF up to 0.4 Hz power msec<sup>2</sup> | 21.25 (14.56-42.88) | 27.32 (9.36-92) | 0.798 |
|  | LF/HF up to 0.4 Hz ratio | 1.98 (0.71-2.31) | 2.79 (1.71-4.96) | 0.110 |
|  | Total power up to 0.4 Hz msec<sup>2</sup> | 1165.42 (739.84-2647.93) | 2123.82 (766.91-3701.38) | 0.629 |
|  | VLFn % up to 0.4 Hz % | 93 (91-94) | 90 (82-92) | 0.085 |
|  | ***LFn % up to 0.4 Hz %*** | ***4.5 (3-5.6)*** | ***8 (5-12)*** | ***0.015*** |
|  | HF<sub>n</sub> % up to 0.4 Hz % | 3.1 (1.9-4.1) | 3 (1.5-5) | 0.887 |
| Nonlinear analysis | SD1/SD2 ratio % | 21 (17.5-35) | 26 (14.4-35) | 0.887 |
| | DFA α1 | 1.12 (0.75-1.15) | 1.12 (0.89-1.37) | 0.376 |
| | DFA α2 | 1.23 (1-1.3) | 1.22 (1.09-1.26) | 0.842 |
|  | Correlation dimension D1 | 1.71 (1.45-2.51) | 1.68 (1.28-2.3) | 0.670 |

Values are expressed as median and interquartile range. DFA: detrended fluctuation analysis; D1 correlation dimension. In bold and italics, $p<0.05$ . VLF: very low frequency power; LF: low frequency power; HF: high frequency power; VLFn%: very low frequency normalized power, LFn%: low frequency normalized power; HF<sub>n</sub> %: high frequency normalized power; DFA $\alpha$ 1: short term 4-16 beats fractal correlation; DFA $\alpha$ 2: long term 16-64 beats fractal correlation; SD1: HRV short-term variability; SD2: HRV long-term variability.

In the frequency domain analysis, power absolute values were not different. Although LF power was also lower (28.62 vs 70.43 msec), it did not reach statistical significance (p=0.097). However, when comparing normalized values, patients who died at the 28-day had a lower median LFn% than those who survived, 4.5% vs 8% (p=0.015). When performing a ROC curve for 28-day mortality of LFn% we found an area 0.825 (95% CI: 0.66-0.98, p=0.017). An optimal LFn% cut-off (Youden’s index) was 5.27% for a sensitivity of 83.3% and a specificity of 76.2% (Fig 2). VLFn% was slightly higher in non-survivors, although not reaching statistical significance (93 vs 90 %, p=0.085).

**Fig 2.**
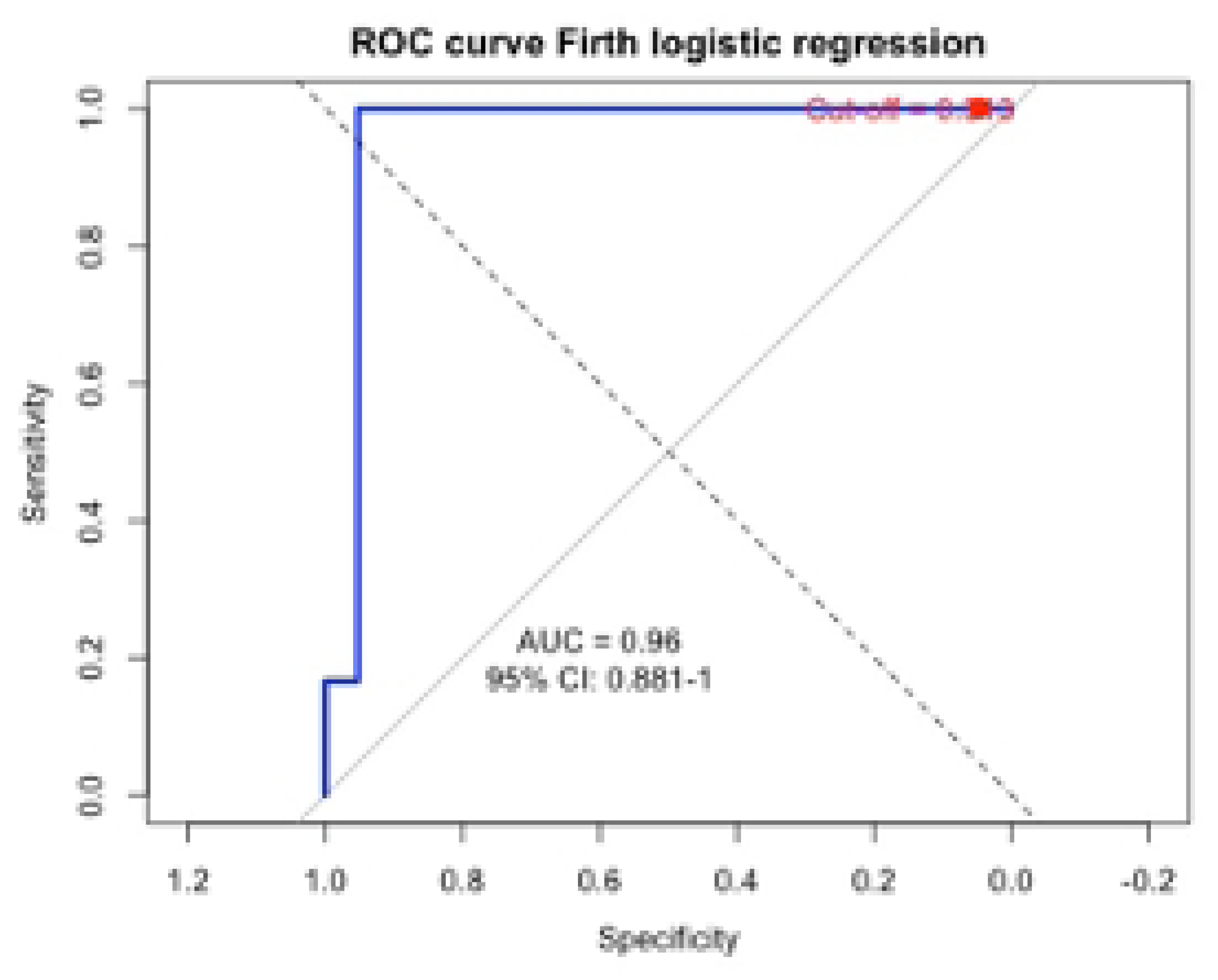
ROC curve for 28-day mortality of LFn%.

There were no differences using non linear analysis variables.

### BINOMIAL LOGISTIC REGRESSION MODEL FOR 28-DAY MORTALITY

We performed a univariate analysis using Firth’s logistic regression and found that uremia, procalcitonin, age and the use of RAASi were related to 28-day mortality. Concerning HRV, the Firth’s logistic regression model using LFn% at 0.4 Hz was statistically significant (χ² = 5.71, p = 0.017). LFn% at 0.6 Hz was also significant (χ² = 5.79, p = 0.016). VLFn% had also a positive association although not reaching statistical significance (χ² = 2.91, p = 0.088) (table 3).

**Table 3.** Univariate binomial robust logistic regression – 28-day mortality.

|  | Beta | Standard error | fd | Sig. | OR | 95% C.I. for OR |  |
| --- | --- | --- | --- | --- | --- | --- | --- |
|  |  |  |  |  |  | Inferior | Superior |
| Age | 0.0755 | 0.048 | 1 | 0.048 | 1.078 | 1 | 1.285 |
| APACHE II score at admission | 0.162 | 0.094 | 1 | 0.068 | 1.173 | 0.989 | 1.478 |
| RAASi | 2.657 | 1.062 | 1 | 0.005 | 14.15 | 2.127 | 168.848 |
| Urea | 0.093 | 0.038 | 1 | 0.004 | 1.097 | 1.026 | 1.211 |
| Procalcitonin | 2.174 | 1.104 | 1 | 0.031 | 8.793 | 1.212 | 177.327 |
| LFn% at 0.4 Hz | -0.383 | 0.195 | 1 | 0.017 | 0.682 | 0.388 | 0.946 |
| VLFn% at 0.4 Hz | 0.16 | 0.11 | 1 | 0.088 | 1.18 | 0.982 | 1.722 |

*RAASi: Renin-angiotensin-aldosterone system inhibitors; LFn%: low frequency normalized power, VLFn%: very low frequency normalized power*

We performed a multivariate Firth’s logistic regression using LFn% at 0.4 Hz adjusted to RAASi use, urea and procalcitonin as covariates. Concerning RAASi use, the model was statistically significant (likelihood ratio test: χ² = 9.04, df = 2, p = 0.0109) but neither LFn% at 0.4 Hz (OR= 0.73, 95% CI 0.33 to 1.113, p = 0.159) nor RAAS inhibitor use (OR= 6.031, 95% CI 0.85 to 70.81, p = 0.073) reached statistical significance individually after penalization. After adjusting using procalcitonin, the model was statistically significant (likelihood ratio test: χ² = 7.78 df = 2, p = 0.020) but, again, neither LFn% at 0.4 Hz (OR= 0.75, 95% 95% CI 0.45 to 1.018, p = 0.068) nor procalcitonin (OR= 5.15, 95% CI 0.55 to 486.871, p = 0.16) reached statistical significance individually.

Finally, when adjusting LFn% at 0.4 Hz with urea as a covariate, the model was statistically significant (likelihood ratio test: χ² = 12.20, df = 2, p = 0.0022). After penalization, LFn% at 0.4 Hz and urea remained statistically significant.

We performed a ROC curve for the multivariate model. When performing a ROC curve, we found an area of 0.96 (95% CI: 0.88-1, p=0.017). Optimal LFn% cut-off was 0.313 (Youden’s index) with a sensitivity of 100 % and a specificity of 95.2% (figure 3).

**Figure 3.**
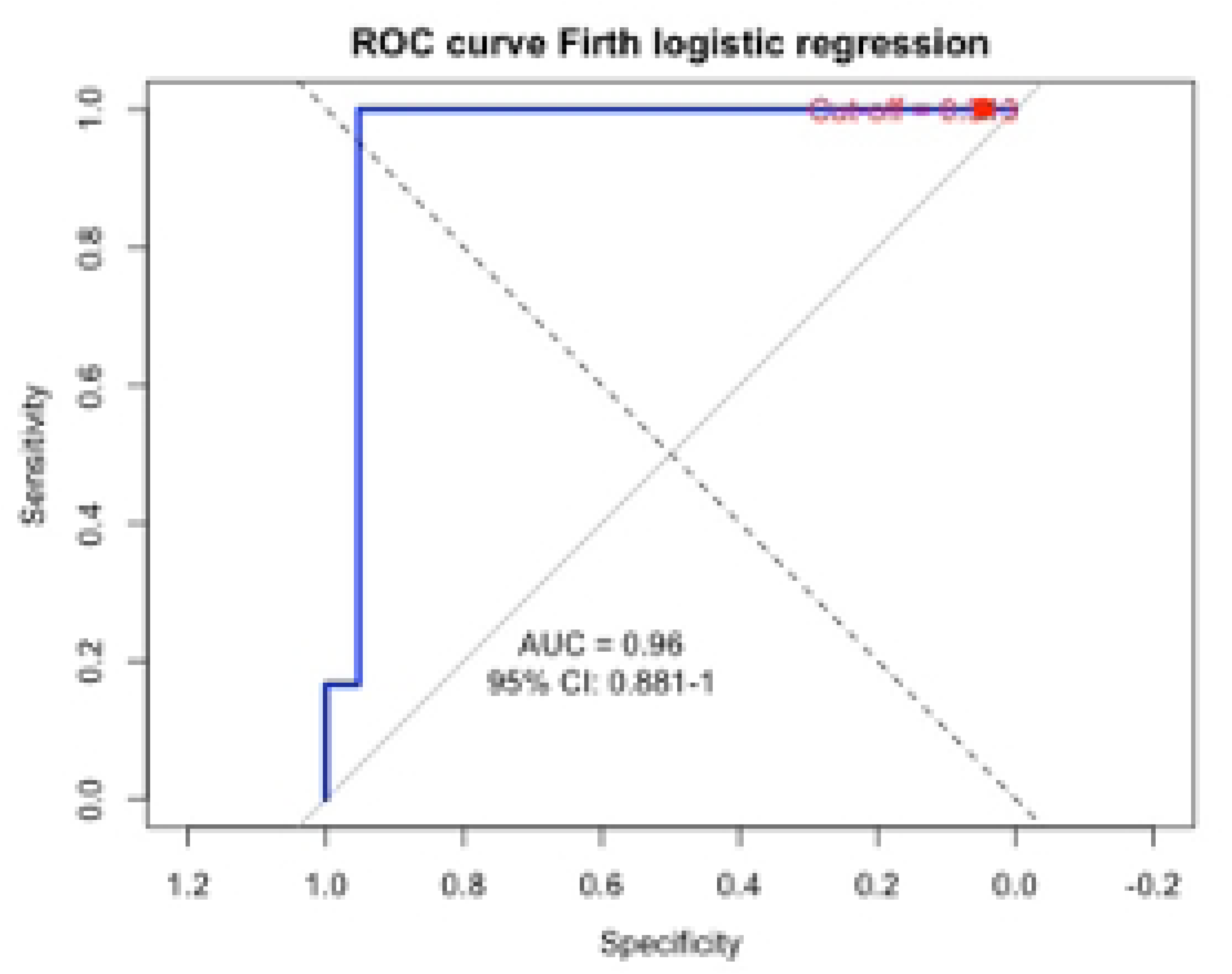
ROC curve multivariate Firth’s logistic model for 28-day mortality using LFn% and urea

We found no association of HRV variables with IMV need.

### LINEAR REGRESSION MODEL FOR ICU LOS

Although not statistically significant, median ICU LOS was longer in non-survivors than in survivors. We performed a linear regression model to analyse the relationship between ICU LOS with other variables at admission. In the univariate analysis, only FiO_2_ and SAF index were correlated with ICU LOS. However, a negative tendency was also found for body temperature, ROX index and DFA α 2.

A multivariate generalized Gamma regression model with log link was constructed, incorporating the SAF index, DFA-α2, and the admission APACHE II score. Although APACHE II is a global severity score, its calculation already integrates several clinically relevant variables—such as age, respiratory rate, and selected biochemical parameters. For this reason, APACHE II was included in the model as a composite indicator, rather than entering its individual components separately.

The standard Gamma regression linear model explained 54.9 % of the variability of ICU LOS (pseudo R^2^=0.549). Only, SAF index was found statistically significant. However, after performing influence diagnostics, a very high Cook’s distance (0.27) was found for observation 19. Also, three observations in the predictors had high leverage values (observation 6, 8 and 24 with hat values around 0.34).

Therefore, a robust Gamma linear regression model using M-estimator was fitted. This model explained 41.9 % of the variability of ICU LOS (pseudo R^2^=0.419) explained by a less influence of the outliers; however, Root Mean Square Error was very similar (16.51 vs 16.50, respectively). Cohen’s f^2^ was 0.72, representing a large effect size. Using this model, we found that all predictors were associated with ICU LOS. It must be noted that DFA ⍺ 2 had wide 95% confidence intervals (table 4 and figure 4).

**Figure 4.**
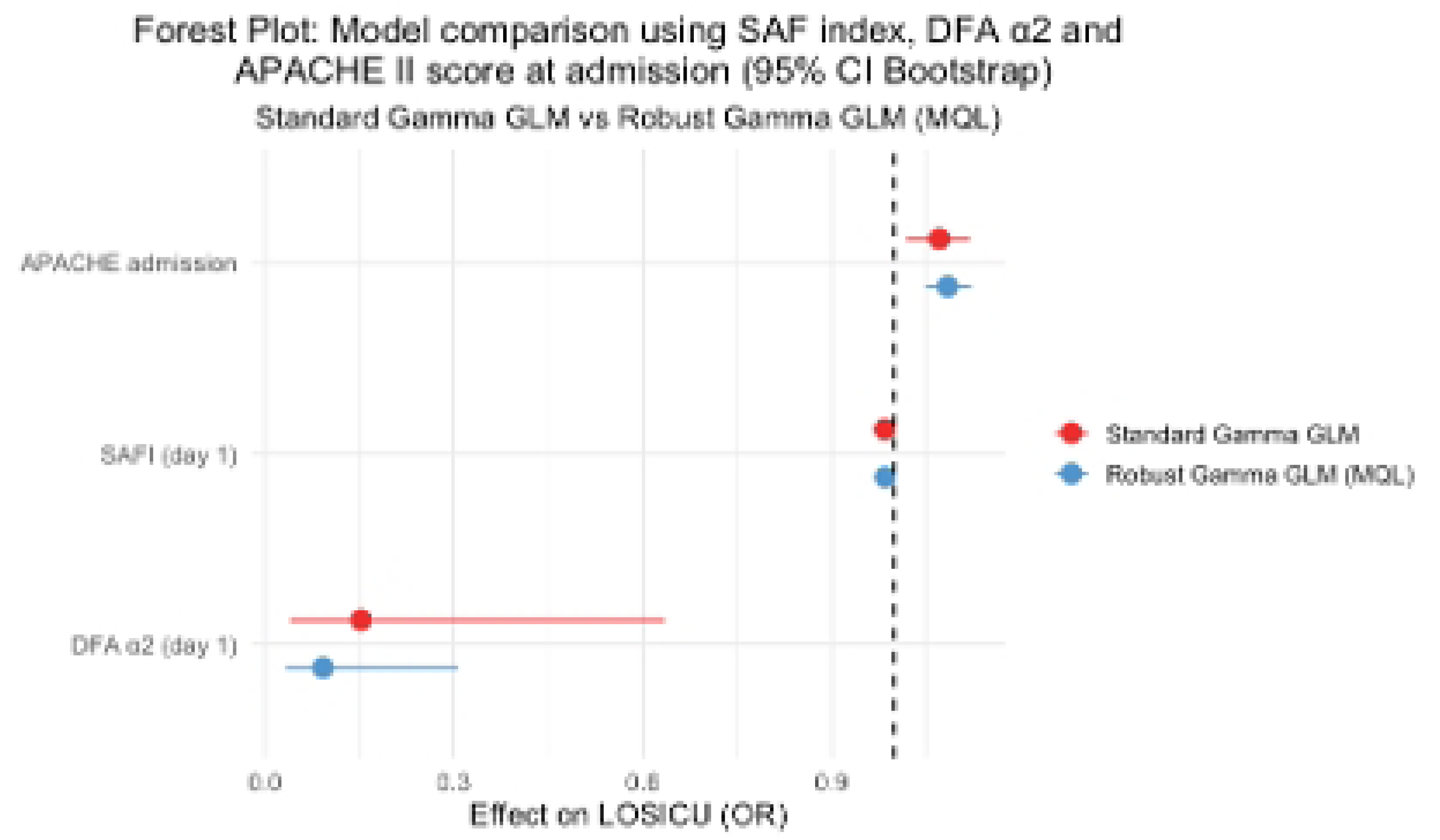
Forest plot: Linear model comparison using SAF index, DFA ⍺ 2 and APACHE II score at admission

**Table 4.** Multivariate linear regression analysis for ICU LOS using APACHE II at admission, DFA ⍺ 2 and SAF index.

|  | Unstandardized<br>coefficients |  | t | Sig.<br>(bootstrapped) | RR<br>(bootstrapped) | 95% C.I. for RR<br>(bootstrapped) |  |
| --- | --- | --- | --- | --- | --- | --- | --- |
|  | Beta | Std. error |  |  |  | Inferior | Superior |
| Generalized Gamma linear model |  |  |  |  |  |  |  |
| APACHE II | 0.069 | 0.036 | 1.903 | 0.017 | 1.071 | 1.020 | 1.123 |
| DFA $\alpha$ 2 | -1.880 | 1.289 | -1.459 | 0.033 | 0.152 | 0.037 | 0.789 |
| SAF index | -0.016 | 0.004 | -3.658 | <0.001 | 0.985 | 0.979 | 0.989 |
| Constant | 6.608 | 1.629 | 4.056 | <0.001 | 741.335 | 150.48 | 3707.42 |
| Robust generalized Gamma linear model |  |  |  |  |  |  |  |
| APACHE II | 0.081 | 0.013 | 6.07 | 0.002 | 1.084 | 1.046 | 1.123 |
| DFA $\alpha$ 2 | -2.38 | 0.476 | -5.01 | 0.003 | 0.092 | 0.031 | 0.361 |
| SAF index | -0.015 | 0.0016 | -9.47 | <0.001 | 0.985 | 0.982 | 0.990 |
| Constant | 6.80 | 0.602 | 11.3 | <0.001 | 899.773 | 195 | 3071 |

The ICU LOS can be expressed by the equation:

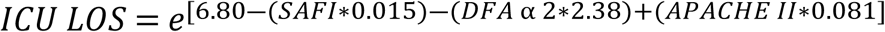

Based on the observed effect size (f² = 0.72), we estimated the sample size needed to achieve a statistical power of 80%. A sample size of approximately 19 patients was calculated (*pwr.f2.test*, {pwr} R package); supporting the adequacy of our sample size to detect the effect.

## DISCUSSION

This is to our knowledge the first prospective study exploring HRV metrics in a sitting position during the first day of ICU stay and its utility to predict mortality in Covid-19 patients. We found that LF normalized power was lower in non-survivors and was associated with 28-day mortality outcome. DFA ⍺ 2 was the only HRV variable associated with ICU LOS.

We developed a statistical model, including DFA α2, admission APACHE II score and SAF index, to explain variation in ICU length of stay in this cohort, using variables that are easy to obtain and compute from medical records within the first 24 hours of ICU admission. Although DFA ⍺ 2 is not routinely measured in clinical practice, we propose that the combination with other clinical data can offer early insights into patient prognosis. The inclusion of DFA ⍺ 2 in future predictive models could be valuable if further studies make this technique easier to compute and analyze in the clinical setting.

### VLF and its relationship to RAAS

In our population, all the patients who did not survive had arterial hypertension and most of them (5/6, 83.3%) were previously treated with RAASi. Arterial hypertension has been described as an important factor associated with severe Covid-19 disease. Several articles describe the relationship between RASS and SARS-CoV-2 (48–50). Initially, RAASi was also associated with severe Covid-19 disease but later publications dismissed this hypothesis (51–53).

The virus interacts with the renin-angiotensin-aldosterone system while using the ACE-2r to enter the cell. This reduced concentration of available enzyme produces a higher concentration of angiotensin II (that has vasoconstrictor and profibrotic functions and, therefore implications in the severe Covid-19 pathophysiology) and lower concentration of angiotensin 1-7. VLF power is increased while using RAASi and is also related to the parasympathetic outflow. Taylor et al. found in ten healthy patients, in supine position, that enalaprilat increased VLF power but did not increase LF and HF powers and that parasympathetic blockade with atropine nearly abolished all power bands including VLF (54). del Valle-Mondragón et al. compared the angiotensin plasma concentration and HRV metrics values during haemodialysis in non-Covid patients. It should be noted that they used the same wavelet approach for HRV analysis as the one applied in our study. These authors found a positive correlation with VLF and LF/HF and a negative correlation with LF and HF power with angiotensin II plasma concentration (55).

Few studies have analysed VLF power in critically ill patients, and many of them show significant methodological problems, such as not reporting the recording duration and the inclusion of sedated or mechanically ventilated patients (47). Chen et al. used HRV metrics to predict in-hospital mortality in septic patients at admission in the emergency department. They described 132 adult patients and found that non-survivors had a diminished HRV than survivors (56). Ten patients did not survive, these non-survivors were hypotensive (mean systolic blood pressure 89 mmHg) and 30% had arterial hypertension or chronic renal disease or diabetes. Compared to our data, we found that our sample had a higher VLFn% but lower LFn% and HFn%. Another interesting issue is that survivors in Chen’s study had higher value of VLFn% but in our study that happened in non-survivors (although not reaching statistical significance, p=0.085) (Supplemental material, table 1). This important difference of HRV powers to VLF bands could be a pathophysiological feature of the Covid-19 disease, as vasomotor tone and blood pressure are maintained in the initial stages of the disease, regardless the outcome (57), in contrast to non-Covid septic patients that usually present in shock and require vasoactive drugs. Another possibility may be methodological issues. Although Chen et al. reported VLF values, they used frequency bands settings for 5-minutes recordings defined below ≤0.04 Hz; we used longer recordings for better spectral resolution within VLF band without the ULF noise and analysed it within 0.003 – 0.04 Hz. The elevated absolute values we report, in comparison to theirs, stem from a more prolonged period of VLF event acquisition. Our recordings were made in the sitting position that also augments VLF band values (58); Taylor et al. showed that a 40° tilt also increased VLF power (54). However, solely this explanation might not be suitable for such large difference.

**Supplementary material. Table 1.**
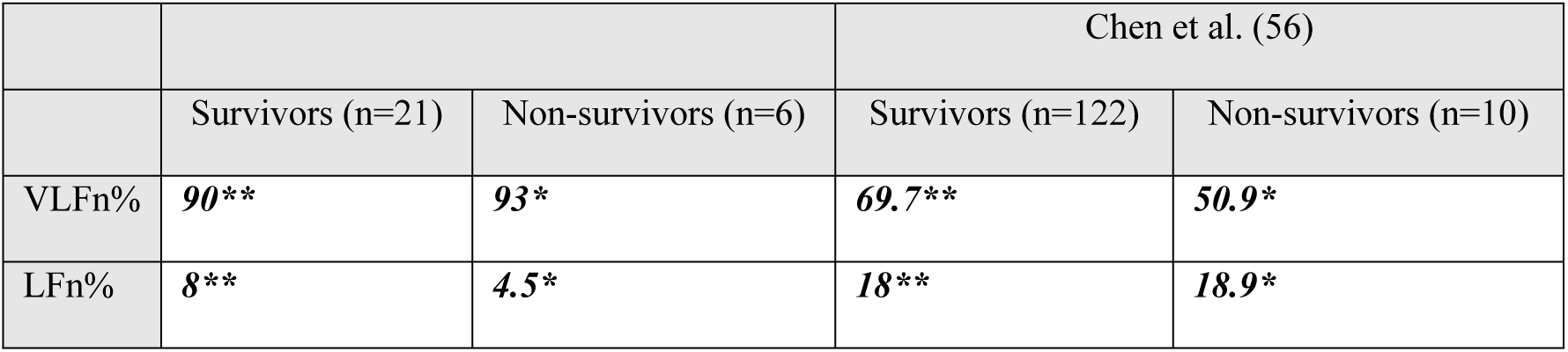
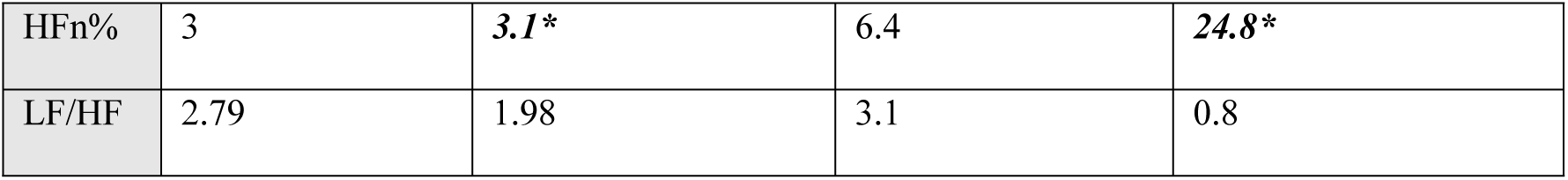
Comparison of normalized HRV metrics with non-Covid septic patients group published Chen et al. In bold and italics, *p<0.05, **p<0.001; one-sample Wilcoxon signed-rank test.

We speculate that the high absolute and relative (VLFn %) values of VLF power found in our study can be related to this Covid-19 pathophysiology where angiotensin II levels are increased. Unfortunately, angiotensin levels or catecholamines were not measured in our patients to confirm this mechanism.

Concerning Covid-19 disease, we found only two studies analysing VLF. Kamaleswaran et al. published a retrospective study of Covid-19 patients admitted in several United States ICUs and compared to an all-cause sepsis database. They found that in the first 8 hours of ICU stay mean VLF and mean LF powers were lower in Covid-19 patients compared to all-cause septic patients (59). However, they did not report whether the patients were ventilated or sedated, nor their body position. Compared to our findings, where VLF absolute power was relatively high (mean 782.15 msec²), the data reported by Kamaleswaran et al. (mean 321.02 ± 9.5 msec²) suggest an overall lower autonomic activity in their cohort during the early ICU period. This discrepancy might be explained by methodological differences, including recording duration, patient positioning (we used sitting position) (60), or the effects of sedation and mechanical ventilation, which were absent in our cohort. Milovanovic et al. analysed cardiovascular reflexes comparing 75 Covid-19 patients at hospital admission against 77 healthy controls: 45 patients had confirmed interstitial pneumonia (severe group) and its mean VLF absolute power was 639.51 ± 3232.01 msec^2^; higher than in mild Covid-19 and control groups but with no statistical significance (61). Compared with our study, the absolute VLF power was similar to that of the severe group (639.51 vs. 782.15 msec^2^, p = 0.052 one-sample Wilcoxon signed-rank test) but higher than that of the control group (500.73 vs. 782.15 msec^2^, p=0.007 one-sample Wilcoxon signed-rank test). Interestingly, both Milovanovic’s and our findings suggest a distinct autonomic pattern in severe Covid-19, possibly reflecting early dysregulation of the sympathetic nervous system or RAAS-related mechanisms. One potential explanation for the higher VLF values observed in our cohort may be the body position during HRV acquisition: while Milovanovic et al. did not specify patient positioning, our measurements were performed with patients in a seated position, which is known to enhance VLF power (60, 62, 63). This methodological difference could partly account for the discrepancies observed between the two studies.

### LF and HF powers

We found that normalized LF power is lower in non-survivors but only if VLF power is considered in the equation (LFn %); if VLF is not considered for normalization (VLFnu, as proposed by the 1996 Task Force), this difference is no longer significant. This finding underscores the importance of taking into account the VLF power band when interpreting HRV measures in our patients.

First, we must consider the methodological issue. As described, we used a wavelet approach that did not slice the sample into windows but rather analyse it as a whole and our recordings were longer than those usually reported in literature, allowing for a better identification of very low frequency events. Therefore, VLF power was higher and LF and HF powers lower.

Nevertheless, our results are concordant with the hypothesis that HRV is diminished in Covid-19 as well as in other diseases. To test this statement, first we compared our sample with the control group published by Kim et al. They presented HRV metrics in 49 healthy patients after sitting five minutes (32). They presented their values in logarithmically transformed values (lnLF and lnHF) so, after transformation of ours, we found that both were lower in our sample (Supplemental material. Table 2):

- For lnLF: 4.25 vs 6.1, p<0.001 in survivors and 3.22 vs 6.1 p=0.028 in non-survivors; one-sample Wilcoxon signed-rank test.
- For lnHF: 3.31 vs 5.6, p<0.001 in survivors and 3.05 vs 5.6 p=0.028 in non-survivors; one-sample Wilcoxon signed-rank test.

Our results are like those found in literature about the relationship between LF and HF powers and organ dysfunction or worse outcomes in critically patients.

In non-Covid patients, Behera et al. analysed HRV in 23 septic patients at ICU admission using 5 minutes and 15 minutes sample windows and found that HF was inversely related to admission SOFA score and this relation was stronger while using larger windows (64). Yien et al. applied spectral analysis of HR and blood pressure in 52 patients admitted in a medical ICU. They described APACHE II score daily and compared survivors (n=25) and non-survivors (n=27). All patients were mechanically ventilated. They found an inverse relation between VLF and LF powers with APACHE II in non-survivors (65). Zhang et al. found that in those patients with acute pancreatitis that did not survive LFnu and LF/HF were lower and HFnu was higher. This pattern was similar in those patients that developed multiple organ disfunction (66).

We also compared our results to those presented by Samsudin et al. that described HRV metrics in septic patients admitted in the emergency department, prior to its admission in the ICU, and therefore, patients who were not under mechanical ventilation or vasopressors. It must be noted that most of the patients were Asian and 30% of the patients were previously treated with betablockers or other antiarrhythmic. They analysed one to six minutes ECG segments and performed time and frequency domain and non-linear analysis to compare patients between 30-day mortality groups. Non-survivors had the same pattern: LFnu was lower and HFnu was higher. When compared with our results, we found that our Covid-19 sample had higher LFnu (73.63% vs 46.5%, p<0.001 in survivors and 66.4% vs 32.9%, p=0.028 in non-survivors; one-sample Wilcoxon signed-rank test) but lower HFnu (26.37% vs 52.8%, p<0.001 in survivors and 33.6% vs 66.4%, p=0.028 in non-survivors; one-sample Wilcoxon signed-rank test) (67).

Aragón-Benedí et al. analysed 112 patients in a multicentre study in five hospitals in Spain, France and Latin America using a commercial device designed to assess the autonomic effect of nociception and analgesia. This device provides two indices: Energy and Analgesia Nociception Index or ANI, surrogates of the SDNN or total HRV and HF, respectively. They found that Energy was lower and ANI was higher in non-survivors and proposed that non-survivors had a higher parasympathetic activity and that HRV might be useful for mortality prediction in Covid-19 critically ill patients (46). Our results agree partially with those presented by these authors as normalized HF power was higher, and total power was lower in non-survivors but without reaching statistical significance. However, it must be considered that patients in Aragón-Benedí’s article were all sedated, under mechanical ventilation and more than 40% in each arm were receiving noradrenaline. These characteristics can, by themselves, challenge their results when trying to explain a Covid-19 direct pathophysiological effect. Our patients were not ventilated and were not being treated with sedatives which might impair sympathetic response and/or enhance parasympathetic flow (68–70). Also, they were recruited in an earlier moment of the severe disease (Aragón-Benedí et al. did not report days from symptoms initiation or pre-ICU admission). Ergo, we think that our results are more related to a more profound sympathetic depletion or a blockage of sympathetic response.

Durmargne et al. arrived at similar conclusions. They compared 24 patients with Covid-19 ARDS with 19 patients with no-Covid ARDS, all under mechanical ventilation and sedated. Mortality was higher in Covid-19 ARDS patients, 46% vs 21 %, although not statistically significant. They found that Covid-19 patients were more bradycardic but HRV metrics suggested an increased vagal tone (higher HF power, SDSD and r-MSSD). However, in contrast with our findings, they found that sympathetic response was also increased, as LF power and LF/HF ratio were higher. Again, a comparison with our work is difficult because the time from diseasès initiation to the EKG acquisition was not informed and patients were sedated. Their conclusion is interesting because they related poor outcomes to the SyNS hyperactivation (71) but it might be flawed because patients were receiving vasopressors, in example noradrenaline, and probably (not reported in the paper) corticosteroids, which might had masked the sympathetic diminished response. Our patients were not with vasopressor therapies and therefore that bias is not present.

This sympathetic disfunction hypothesis can be nourished by our finding that non-survivors had a lower LFn% than survivor. Also, in our study, LFn% at ICU admission had a better discriminative power for 28-day mortality in Covid-19 patients than APACHE II score (figure 41). We found an AUC ROC curve of 0.825 compared to 0.63 published by Vandenbrande et al. (72) or 0.73 by Beigmohammadi et al (73). When compared to total HRV power (Energy) published by Aragón-Benedí et al., our results were also better discriminating 28-day mortality, AUC ROC curve 0.825 vs 0.755 (46).

LFn% was inversely related to 28-day mortality in a robust univariate logistic regression model, as well as, urea and procalcitonin. We also found a statistical trend (p=0.088) for VLFn%. A multivariate logistic model was then performed, including urea and LFn% as variables. LFn% was found significant however the 95% C.I. are wide and its association has to be analysed cautiously. Nevertheless, this model had an excellent predictive capacity for 28-day mortality (sensitivity 100%, specificity 95.2%), better than using LFn% solely or those previously published.

Also, this sympathetic depletion hypothesis can explain the association between the intermediate-long term HRV scaling exponent (DFA ⍺ 2) with longer ICU stay. Literature exploring the utility of DFA mostly involves the short-term scaling exponent (DFA ⍺ 1). However, experimental studies in animals and healthy volunteers found a decrease in DFA ⍺ 2 when a sympathetic blockade is performed (74–77). Annane et al. found that patients with septic shock had a diminished HRV and LF/HF and higher plasma levels of norepinephrine than septic patients but not in shock. They speculated that the high sympathetic drive was impaired via a centrally mediated system (16). It must be noted that the use of betablockers in our population was minimal, therefore, we consider that patients who did not survive were sicker and more sympathetic impaired than those who survived. Additionally, several studies had described the relation between autonomic nervous system, particularly the parasympathetic branch, and inflammation. Papaioannou et al. and Arias-Colinas et al. found that HRV was inversely related with inflammatory biomarkers in patients with an infection (78, 79). In Covid-19 patients, Hasty et al. and the previously commented paper by Mizera et al. found similar results (80, 81). Kevin Tracey described the “inflammatory reflex” as how the nervous system regulates inflammation (82) and Borovikova et al. described the efferent pathway through the nerve vagus. This cholinergic pathway, mediated by the specific α7nAChR receptor, decreased proinflammatory cytokine production in stimulated macrophages (83, 84). In an experimental ventilator-induced lung injury mouse model, dos Santos et al. found that vagus nerve stimulation (electrical and pharmacological) diminished lung inflammation (85).

Although we propose that the diminished LFn% could be related to a Covid-19 pathophysiological characteristic, we also found that LFn% is inversely associated to age, APACHE II score and female sex; which is concordant to the previously published literature (86–88). As previously stated, the lack of age- and gender-matched controls may limit generalisation of our results.

On the other hand, we found no association between SAF index and LFn%. That finding is also interesting as, although the main clinical feature of Covid-19 is respiratory, its effects affect the whole organism. Using only a respiratory parameter to evaluate these patients may be insufficient.

As a conclusion, we propose that more severely ill Covid-19 patients might have a diminished sympathetic response, by depletion of neurotransmitters or by direct blockade, based by the lower LF power and LFn% found in our sample. This idea can explain why severe Covid-19 patients are not tachycardic in the initial stages of the disease despite being hypoxemic. We did not measure serum catecholamines, like the study by Annane et al. (16), to confirm this theory. We cannot assume that the parasympathetic activity via nerve vagus might be enhanced at ICU admission as they were no differences between groups in HF power.

Using HRV metrics, such as LFn% calculation or DFA ⍺ 2 to design diagnostic and prognostic tools, might enhance their predictive and explanatory capabilities.

### ICU LOS and HRV

Although we found no differences in ICU LOS between 28-day mortality groups, we did find that ICU LOS was longer in those patients who required mechanical ventilation. Considering the statistical distribution of the outcome variable, ICU LOS, and the presence of outliers in both outcome and predictors, we performed a standard Gamma and a robust Gamma regression models. The only HRV metric that was related to ICU LOS was DFA α 2.

We propose a linear model that includes APACHE II score, an “organ failure” variable, SAF index, a “therapeutics-related variable” and DFA ⍺ 2, HRV variable. We found that DFA ⍺ 2 and SAF index maintain a negative statistical association and APACHE II score at admission a positive association with ICU LOS (table 4, figure 4).

In our study, the values of DFA α 2 were higher than those published in other populations (37, 89, 90). Beckers et al. described several HRV metrics, particularly nonlinear, between gender groups and how age affects them. They compared 135 women and 141 men, between 18 to 71 years and recordings in day- and night-times. They found that all nonlinear metrics decline with age except DFA α 2 that had a positive correlation (two-tailed Pearson *r* 0.458 for male and 0.604 for female in daytime). They also found that absolute and relative LF power were inversely related to DFA α 2 (91). Other studies have described that both DFA α 1 and DFA α 2 parameters were inversely related to mortality; however, they did not analyse or describe ICU LOS (92–95). On the other hand, Papaioannou et al. described 53 patients admitted in a multidisciplinary ICU and found a positive relation between DFA α 2 and ICU LOS (Pearson coefficient 0.55, p=0.02) (39). In an interesting theoretical article, Francis et al. described that detrended fluctuation analysis and frequency domain metrics are mathematically coupled. They proposed that DFA α2 can be approximated by the relation

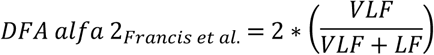

and showed this estimate was highly correlated (Pearson correlation 0.94, p<0.0001) (96) with fractal DFA α 2 as proposed by previous publications (97, 98). Finally, they stated that DFA is not different to frequency domain spectral analysis (96). This latter article might explain why our Covid-19 patients have higher DFA α 2 values than those published. Applying this formula to our data we found that our median value is 1.865 (IQR 1.78-1.90) which agrees to our findings that, in Covid-19 patients, VLF band predominates. Also, this mathematical coupling explains why DFA α 2 in our sample is near 2; as LF power is very small, leading the right-hand side of the equation close to one, therefore 2*1=2. Another important issue that must be analysed is that Francis et al. used autoregressive spectral methods which might underestimate power spectrum compared to a wavelet approach (43).

It must be noted that VLF nor LF were associated with ICU LOS. This situation might be related to the very structure of DFA. DFA measures the entire variability of the signal rather than the power (variability) within a specific frequency band. ICU LOS is a complex continuous outcome; it depends on many factors: disease-related, therapeutics, background and cannot be attributed to a single organ or system. Following this reasoning, DFA may be a more appropriate metric for describing ICU LOS as, both, measure complex, non-linear relationships within every patient’s evolution. This proposal can be nourished with the finding that between 28-day mortality groups, there are no differences in DFA ⍺ 2 values (p=0.86, Mann Whitney U test) but, when using 90-day mortality as an outcome, we observed a statistical trend (p=0.11, Mann Whitney U test). This change was not observed in any other HRV metric.

Khodadadi et al. published a population-based cohort study, describing 41 Covid-19 patients and found a weak inverse relationship between LF power and ICU LOS (Pearson *r* = −0.53, 95% CI: −0.73 to−0.24) (99). Prabhakar et al. published a retrospective study describing 343 septic patients and found that LF/HF ratio (2.8 vs. 1.5, p = 0.02) and DFA α 2 (1.0 vs. 0.7, p < 0.01) were lower in non-survivors at 30^th^ day. Also, LFnu was lower and HFnu higher in non-survivors; absolute VLF was not different (170 vs 216 msec^2^, p=0.53). They propose a logistic model combining qSOFA score and DFA ⍺ 2 which was better than qSOFA alone and qSOFA + SDNN to predict 30-day mortality (AUC 0.76 vs 0.68 vs 0.71). Interestingly, adding LF/HF ratio to the qSOFA+DFA ⍺ 2 model did not increase AUC (94). These results strengthen our findings and the proposal that DFA ⍺ 2 and LFn% may provide prognostic information regarding the systemic impact of Covid-19 disease.

## CONCLUSIONS

We found that LFn% is lower in non-survivors Covid-19 patients and proposed that this finding might be related to a diminished sympathetic response in early phases of severe disease. Also, this is the first study to analyze HRV in critically ill patients in the sitting position, regardless of their previous diagnosis. We found that DFA ⍺ 2 was related to longer ICU LOS and describe a simple model to predict ICU length of stay in Covid-19 patients that could be further explored for clinical predictive and early warning models in a more extended population. We also propose that, when analyzing HRV in critical illness, records should be longer than 5 minutes to capture VLF events. This information might be prognostic and physiopathologically relevant. Our results are consistent with previously published articles that supported the hypothesis that the cardiovascular response in COVID-19 is centrally mediated. Validation in a larger and more balanced dataset is recommended.

## Data Availability

All relevant data are within the manuscript and its Supporting Information files.

## ACKNOWLEDGMENTS

the present affiliation of Tomás Fariña-González is the Department of Intensive Care Medicine, Hospital Universitario Infanta Sofia.

## STATEMENTS AND DECLARATIONS

### Availability of data and materials

The datasets used and/or analyzed during the current study are available from the corresponding author on reasonable request.

### Author contributions

TFF, FMS and MEH contributed to the study conception and design. TFF, FMS, MEH, JL and IO contributed with material preparation, data collection and curation. Analysis was performed by TFF and FMS. The first draft of the manuscript was written by TFF and all authors commented on previous versions of the manuscript. All authors read and approved the final manuscript.

### Ethics approval

The study protocol was approved by the Hospital Universitario Clínico San Carlos Ethics committee (CI 21/169.E).

### Consent to participate

Written informed consent was obtained from all patients or their next-of-kin / guardians.

### Consent for publication

not applicable.

